# Microbial Communities Facilitate Pathogen Persistence in Hospital Environments

**DOI:** 10.1101/2025.10.28.25339026

**Authors:** Pratyay Sengupta, Vijay Suryakant Kapse, Karthik Raman

## Abstract

The microbiome of hospital environments is increasingly recognised as a reservoir for clinically relevant pathogens. Yet, the ecological mechanisms that shape pathogen persistence are poorly understood. Here, we ask whether the microbial community structure influences the persistence of the pathogen and if it metabolically supports their proliferation in the harsh conditions of hospitals. We analysed shotgun metagenomic data from a hospital and other urban built environments to explore and compare their microbial diversity. By building and analysing microbial co-occurrence networks across these environments, we show that hospital communities harbour unique microbial interactions, dominated by phylogenetically and functionally diverse keystone pathogens. Combining metabolic modelling with microbial associations, we demonstrate that the hospital microbial communities provided significantly higher metabolic support to pathogens relative to other environments, and quantify it by means of a Pathogen Support Index. This computational framework advances our understanding of pathogen ecology in urban microbiomes, suggesting potential avenues for intervention to mitigate infection risks in healthcare settings.

## Introduction

In the post-industrial era, rapid urbanisation has led to increased habitation in built environments. Microorganisms present in these environments are known to profoundly influence human health by directly altering physiological, immunological, and mental development [1]. Each built environment has a distinct microbiome, and the microbes that populate these environments interact amongst themselves as well as with the human occupants in complex ways [2]. Within built environments, the microbiome of hospital environments is of particular concern, since hospitals host immunocompromised patients with a high risk of acquiring Healthcare-associated Infections (HAIs) [3, 4]. Previously, attempts have been made to characterise hospital microbiomes using both culture-omics and culture-independent (16S amplicon and shotgun metagenomics) methods [5, 6, 7, 8]. Many of these studies have focused on surface microbiomes, while a few have characterised hospital microbiomes through wastewater sampling [9, 10]. However, these findings may be confounded by the advanced treatment processes that could eliminate bacterial contamination, potentially obscuring the microbiome signatures of the hospital [11]. Air sampling methods have also been used to estimate the impact of microorganisms carried in the atmosphere of hospital settings [12, 13]. The principal objectives of these studies have been to characterise antimicrobial resistance (AMR) profiles and catalogue microbial diversity, with an emphasis on ESKAPE pathogens [14, 15]. Despite these efforts, the underlying microbial interactions that drive community assembly and stability remain hitherto underexplored. Deciphering these interactions between community members is integral to explaining the ecological processes related to community stability, pathogen colonisation, and microbiome resilience within hospitals and other buildings.

In microbial ecosystems, species interactions are typically driven by metabolic dependencies [16]. Species co-occurrence itself has been shown to produce metabolic interdependence [17]. Co-occurrence network analysis remains among the most widely used methods in assessing the ecological relationships of complex microbial communities [18, 19]. Although co-occurrence networks offer valuable insights into microbial association patterns, they are fundamentally not suited to distinguish real biotic interactions [20]. In addition, recent advances in sequencing technologies and microbial genomics have paved the way for metabolic modelling approaches that go beyond correlating associations—to inferring ecological mechanisms such as nutrient exchange, metabolic cooperation, and competition [21, 22]. Recently, large-scale community metabolic modelling has been leveraged to gain novel insights into microbial community assembly and molecular interaction mechanisms in soils [23], oceans [20], and gut microbiomes [24]; however, to the best of our knowledge, no attempts have been made so far to model urban built environment microbiomes. In this study, we utilised the microbial composition of 438 hospital samples from Tan Tock Seng Hospital, a major tertiary-care facility and national referral centre for communicable diseases in Singapore, which was previously quantified by Chng *et al.* through shotgun metagenomic sequencing [6]. To contrast hospital microbiomes with other urban environments, we included metagenomic data from an indoor office environment at the Genome Institute of Singapore [6], where the sampling sites were selected to represent locations similar to those found in the hospital, and from frequently touched surfaces at metro stations across Singapore [25]. This comprehensive dataset enabled us to explore microbial diversity, species interactions, community structure, and the role of pathogens in the shaping of microbial ecosystems — areas that have remained largely unexplored in previous studies.

Here, we present an integrated framework that combines network-based analysis and genome-scale metabolic modelling to dissect microbial interactions and their functional potential in diverse built environments (Figure 1). First, we compared the microbial diversity in a hospital environment with two other representative urban built environments: a metro station (public transport) and an office space (indoor workplace), all sampled within Singapore. Next, we employed microbial association network analysis to infer robust co-occurrence patterns and identify ecologically relevant microbial associations across these environments. Third, we investigated the metabolic capacity of interacting communities through genome-scale metabolic models that reveal potential metabolic cross-talk and resource sharing among taxa. Finally, to understand whether these microbial communities could support the persistence of pathogens, we developed a pathogen support index to quantify the capacity of specific consortia to host and potentially propagate pathogenic species within these ecosystems. Together, this approach provides a system-level perspective on how the structure, interactions, and metabolic capacity of the microbial community influence the stability and pathogenic potential of the microbiomes of the built environment. These insights could inform targeted interventions for infection control and healthier urban design.

**Figure 1:**
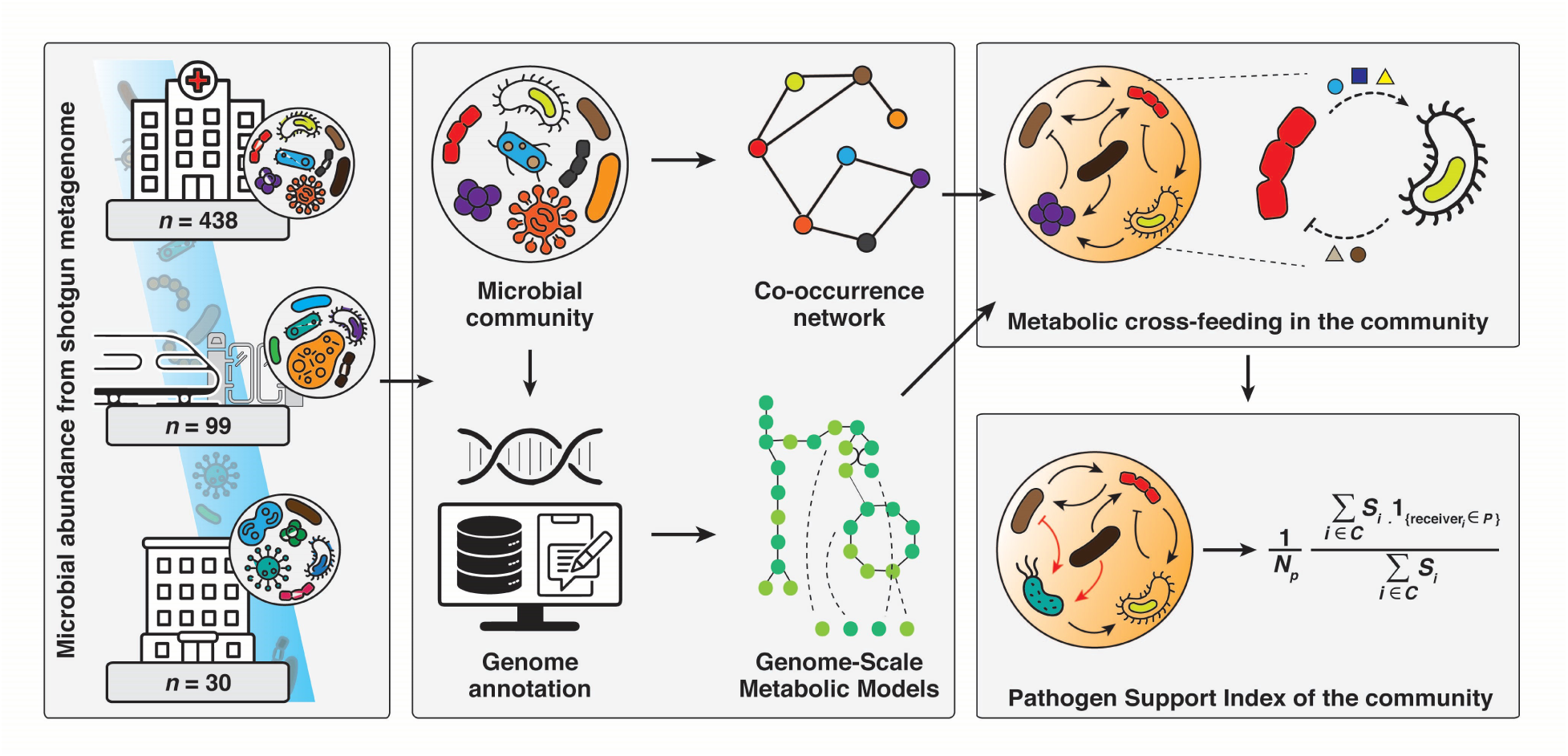
Overview of the study. This figure displays the sources of the microbial community data and their subsequent processing using integrated network and genome-scale metabolic modelling approaches. Utilising these techniques, metabolic cross-feeding and microbial interactions across built environments were investigated. It also highlights the role of the microbial community in supporting and facilitating pathogen colonisation.

## Results

### Microbial communities in hospitals are distinct and diverse from other built environments

The relative abundances of 900 distinct microbial species were estimated from 567 environmental samples spanning three biomes: hospital environments, metro stations, and indoor office spaces [6]. We observed that hospital environments had the highest species richness, with 782 taxa, followed by metro stations (380 species) and office environments (186 species). Regardless of these differences in terms of the actual species counts, bacterial diversity contributed to the majority of the overall microbial abundance for all three biomes (Figure 2a). Notably, 127 viruses were detected in hospital environments, resulting in 9.65% of total abundance. Further, to ascertain the role of the sample size in microbial diversity, species accumulation curves were plotted for each biome. The accumulation curves for the metro and office environments, even when normalised for sample counts, reached a plateau early compared to hospital samples. This indicates that the microbial richness in the hospital environment remains substantial, independent of sample size (Supplementary Figure 1a).

**Figure 2:**
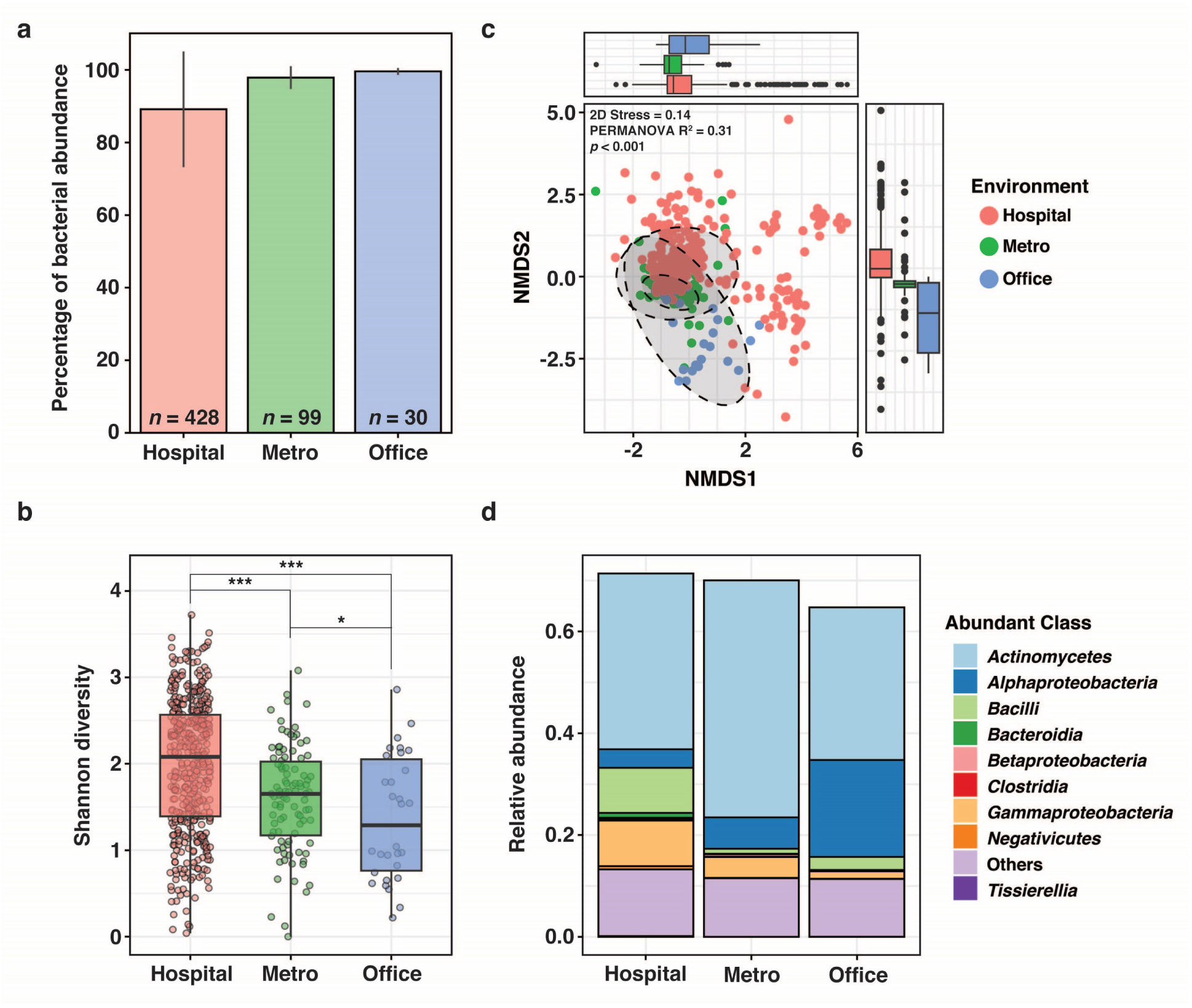
Microbial diversity across the built environments. **a,** Relative abundance of bacterial taxa in samples from hospital, metro, and office environments. Bars indicate mean ± s.d. **b,** Shannon diversity indices across samples. Pairwise comparisons were performed using the Kolmogorov-Smirnov test. Significance levels: *** *p* ≤ 10^−3^, * 10^−2^ *< p* ≤ 0.05. **c,** Beta diversity across environments based on Bray-Curtis dissimilarity, visualised using non-metric multidimensional scaling (NMDS). Statistical significance was assessed using PERMANOVA. Box plots in **b** and **c** show the median, interquartile range (IQR), and whiskers extending to 1.5× IQR; **d,** Class-level taxonomic profiles after filtering for taxa with ≥ 0.1% relative abundance and ≥ 10% prevalence per environment.

Further, to estimate and compare the diversity of microbial communities from the hospital with other studied biomes, Shannon diversity was computed. A significant variation in alpha diversity was observed between environments (Pairwise Kolmogorov–Smirnov test, *p <* 0.01) (Figure 2b). In order to avoid sampling biases due to unequal sample sizes, 100 iterations of rarefied subsampling were performed, with 30 samples per biome randomly selected for each iteration. The results consistently showed that hospitals had the highest microbial diversity, followed by metro stations, while offices had the lowest (Supplementary Figure 1b). In addition to alpha diversity, beta diversity analysis was performed to interpret compositional variation between biomes. Although the samples exhibited overlap along the NMDS1, greater differentiation was observed along the second axis (NMDS2), separating hospital and office samples. The communities of the metro stations showed patterns that overlapped with both hospital and office samples. Moreover, the hospital samples were found to be widely scattered, indicating increased internal heterogeneity (Figure 2c). A PERMANOVA analysis demonstrated significant differences in the composition of the microbial community between environments (*p <* 0.001). Of the variables tested, the sampling site was the most important driver of community variation (*R*^2^ = 0.15, *p* = 0.001), followed by surface material (*R*^2^ = 0.06, *p* = 0.001) and environmental conditions (*R*^2^ = 0.04, *p* = 0.001).

At the class level, the microbial composition also varied considerably between biomes (Figure 2d). Particularly, the hospital environments were enriched with taxa known to be associated with nosocomial pathogens, such as Bacilli, Clostridia and Gammaproteobacteria. In contrast, Alphaproteobacteria were more abundant in office samples. Although common in every biome, Actinomycetes were especially abundant in samples from metro stations. These findings suggest that the environmental context significantly influences the diversity and composition of the microbial population. We noted that hospital environments host not only the most diverse microbial communities but also distinct taxonomic profiles, with a higher viral and pathogen-associated diversity. Metro stations represent an intermediate biome with moderate overlap, while office environments have lower diversity and more uniform community structures.

### Network analysis reveals habitat-specific microbial structuring across urban environments

Next, to understand the microbial interactions within environments, we constructed species-level co-occurrence networks for each environment (Figure 3a). The networks were generated under stringent conditions to reduce noise and minimise false positives (see Methods). Despite such strict selection criteria, these networks captured more than 50% of total bacterial abundance in any biome, widely representing the community structure (Supplementary Figure 2a). The network properties were then assessed using several key topological parameters, such as the number of nodes, edges, and other centrality measures (Table 1). The hospital network emerged as the largest, comprising 83 nodes and 506 edges, followed by the metro station network (43 nodes and 97 edges) and the office network (14 nodes and 30 edges) (Supplementary Table 1). Moreover, the number of nodes and edges in each network was observed to be scaled with microbial richness, indicating the rationale that environmental diversity drives the complexity of the networks (Supplementary Figure 2b).

**Figure 3:**
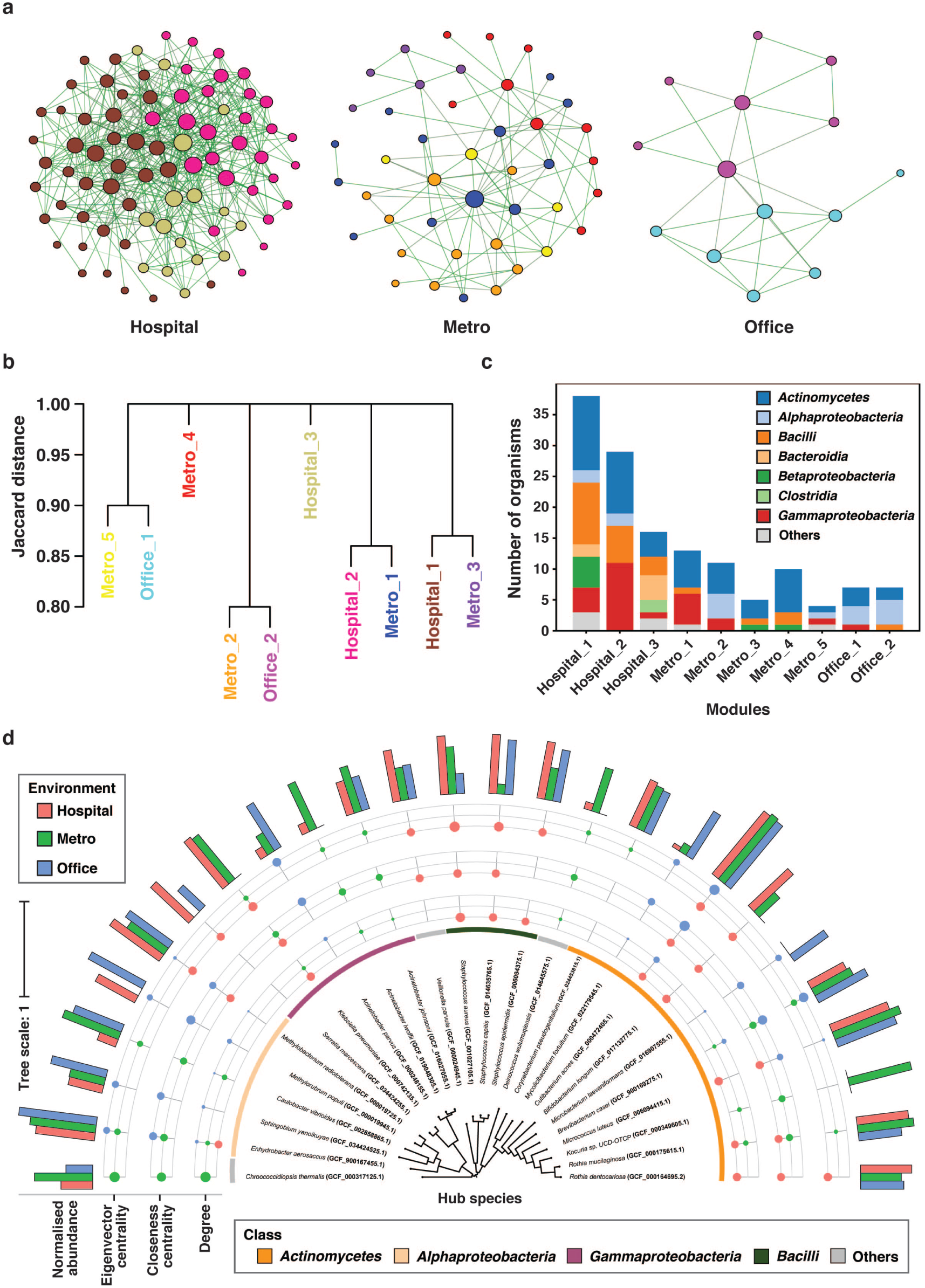
Microbial co-occurrence network analysis. **a,** Co-occurrence networks for hospital, metro, and office environments, where nodes represent microbial taxa and edges indicate significant associations between them. Node size corresponds to normalised degree centrality, and nodes are coloured by network module. **b,** Network modules were hierarchically clustered using Jaccard distance based on shared taxa. Module colours are consistent with those shown in the networks in **a**. **c,** Taxonomic distribution of microorganisms within each module, shown at the class level. **d,** A phylogenetic tree of hub taxa identified across all environments was generated using a set of representative marker genes (see ‘Methods’ section). The inner ring indicates the taxonomic class of the hubs. The middle rings represent the normalised centrality scores (degree, closeness, and eigenvector centralities), where dot size reflects the magnitude and colour denotes the environment. The outer bar plot shows normalised abundance of each hub taxon across environments.

**Table 1:**
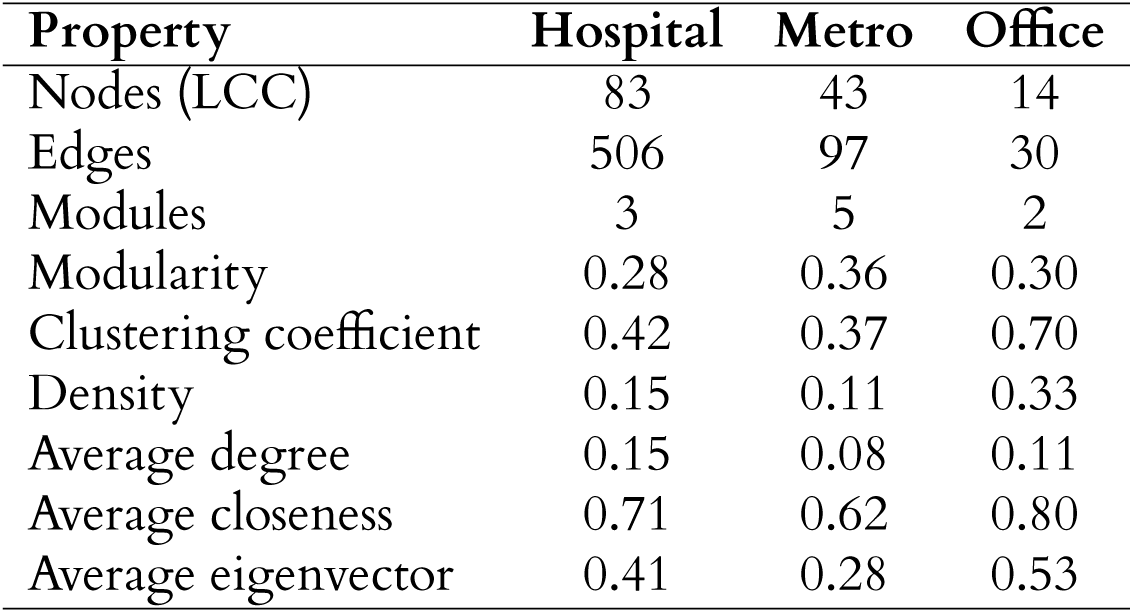
Summary of key topological properties computed from microbial co-occurrence networks in three built environments. LCC stands for the largest connected component.

Further, exploring the modules in the biome networks revealed habitat-specific structures. Here, modules represent densely connected subgroups of taxa, potentially indicating functional or ecological partitioning in networks. We noticed that the metro station network exhibited comparatively higher modularity (0.364), separated into five distinct modules, compared to three in hospitals and two in office networks. Interestingly, the intermodular clustering showed overlap between some hospital and metro modules, as well as between office and metro modules (Figure 3b). This suggests that metro stations could serve as microbial intermediaries between more static built environments. We also summarised the taxonomic distribution of each module to understand their class-level diversity (Supplementary Table 1), revealing distinct patterns and highlighting the variation present between different modules (Figure 3c).

Following this, we identified ten hubs per network with the highest centralities, representing potential keystone taxa in that environment (Supplementary Table 1). A total of 26 hub nodes were selected, distributed primarily in three major classes – Actinomycetes (10), Alphaproteobacteria (5), and Gammaproteobacteria (5) (Figure 3d). In the hospital network, pathogenic species such as *Klebsiella pneumoniae*, *Staphylococcus aureus*, *S. epidermidis*, *S. capitis*, and *Serratia marcescens* dominated the centralities of the network. Additionally, *Rothia dentocariosa* and *R. mucilaginosa* were identified as hubs across hospital networks, with the latter acting as a key node in the metro network as well. Notably, *Enhydrobacter aerosaccus* was the only species that was consistently found to be a community hub in all three habitats. In contrast, the metro station networks featured *Acinetobacter johnsonii*, *A. lwoffii*, *Caulobacter vibrioides*, *Chroococcidiopsis thermalis*, *Deinococcus wulumuqiensis*, and *Kocuria* sp. UCD-OTCP as hubs. Unique hubs like *Acinetobacter parvus*, *Methylobacterium populi*, *Microbacterium laevaniformans*, *Mycolicibacterium fortuitum*, and *Sphingobium yanoikuyae* shaped the office network. Interestingly, *Bifidobacterium longum*, a gut-associated probiotic, appeared as a hub only in hospital settings. Our analysis emphasised the role of the environment in the structure of the microbial community with distinct hub species and phylogenetic diversity. This prompted us to investigate deeper into the microbial interactions and the factors governing interspecies interactions.

### Co-occurrence network identifies specialised microbial associations

We evaluated microbial interactions by exploring the network edges that represent co-occurring associations. We observed that each biome exhibited a large proportion of unique nodes as well as interactions (Figure 4a). The hospital network contained the highest number of unique edges, accounting for 79.6% (485 out of 609), while the metro and office networks had fewer but significant unique interactions – 70.1% and 56.6%, respectively. Surprisingly, all three biomes shared one single interaction, that between *Brevibacterium casei* and *Enhydrobacter aerosaccus*. To further characterise these patterns, edges were classified as either generalised (occurring in multiple biomes) or specialised (unique to a single biome) based on their occurrence across biomes (Figure 4b). The generalised interactions were relatively low in fraction in all the environments, ranging from 4.1% to 23.3%. However, the specialised edges, unique to the biomes, were more prevalent in all networks. We further subdivided these specialised interactions into three types (see Methods):

1. Interactions between two unique nodes were most frequently observed in the hospital environment (18.7%) and least common in the office setting (6.6%);
2. Interactions linking one generalist node with a unique partner were especially prominent in the office, where they accounted for 60% of interactions, followed by 45% in the hospital and 38% in metro stations; and
3. Unique edges between common nodes made up ∼ 30% of edges in both the hospital and metro networks, but only 10% in the office (Figure 4c).

**Figure 4:**
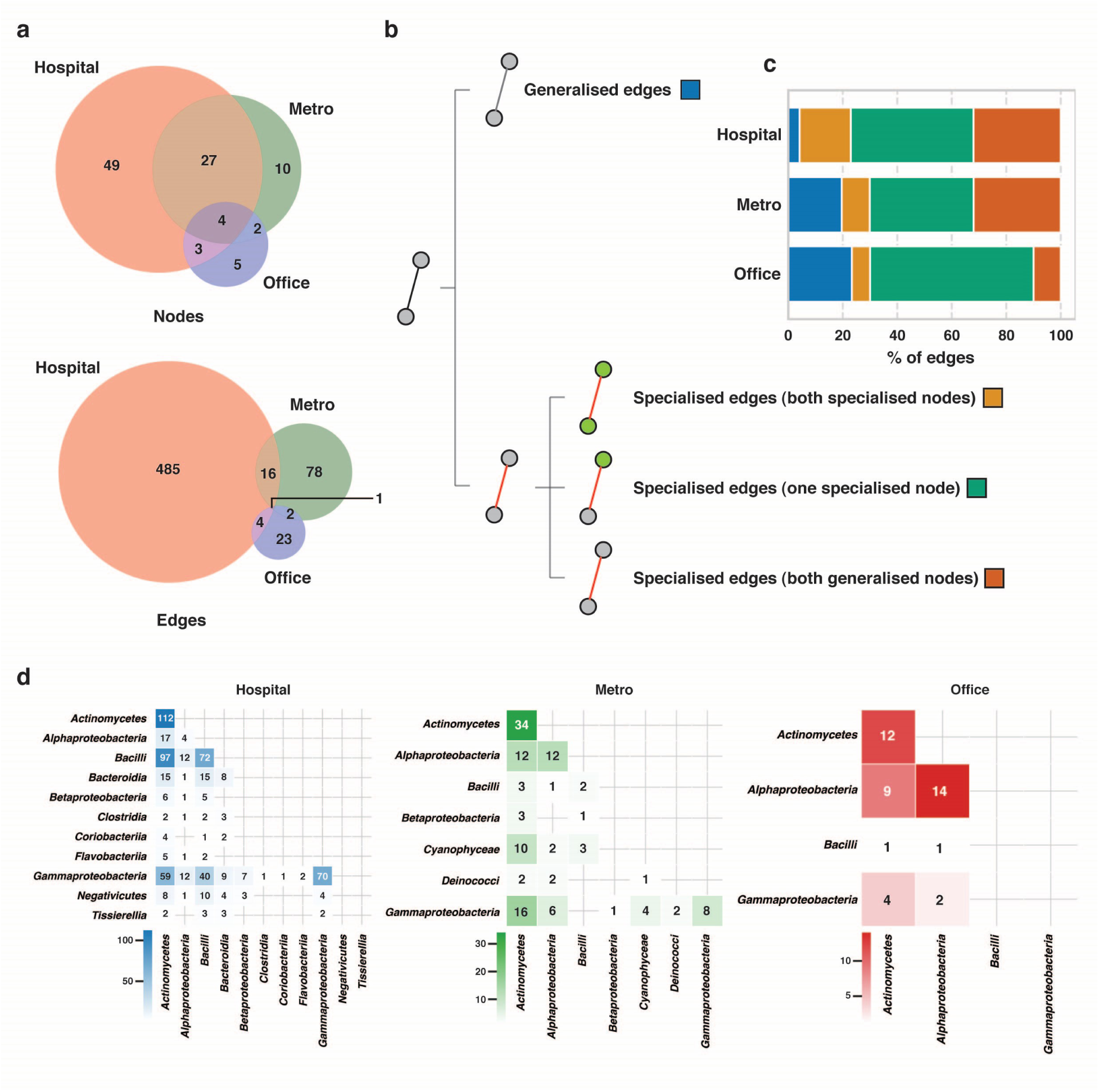
Characterisation of microbial associations through edge analysis. **a,** Venn diagrams showing the overlap of nodes and edges among microbial co-occurrence networks from hospital, metro, and office environments. **b,** Schematic representation of edge classification based on the distribution of interacting taxa across environments. Edges were categorised as generalised (shared across all environments) or as one of three types of specialised edges (unique or shared between subsets of environments; see Methods). Colours boxes next to the categories correspond to those used in **c**. **c,** Distribution of edge categories within each environment, shown as a proportion of total observed edges. **d,** Heatmaps of class–class co-occurrence patterns for each environment displayed interaction strength between microbial classes. Each heatmap uses an individual colour scale with annotated values for all class–class pairs.

This suggests that microbial interactions are contingent upon environmental context, even when the same taxa are present across different settings. Our analysis showed a distinct biome-specific architecture even at the class level (Figure 4d). In hospitals, *Actinomycetes* dominated the interaction network, engaging broadly within their own class along with Bacilli and Gammaproteobacteria, both of which had strong intra-class connectivity. The networks of metro stations were similarly centred around *Actinomycetes*, with additional inter-class interactions involving Alphaproteobacteria and Gammaproteobacteria. In contrast, office environments were structured primarily by interactions between Actinomycetes and Alphaproteobacteria, while Bacilli and Gammaproteobacteria engaged predominantly in inter-class associations.

We subsequently looked at whether class-level patterns contribute to any of the specific edge categories (Supplementary Figure 3). Generalised edges were driven primarily through intra-class interactions, implying common functional roles across environments. In contrast, the specialised edges showed more inter-class interactions. The specialised edges that included two unique nodes were driven largely by *Actinomycetes*, *Bacilli*, and *Gammaproteobacteria*. For edges that joined a generalist and a specialised node, the main contributors were *Actinomycetes*, *Bacteroidia*, *Clostridia*, and *Gammaproteobacteria*. Furthermore, specialised edges involving two generalist nodes featured similar dominant classes as the generalised edges, but displayed a higher proportion of inter-class interactions, with *Negativicutes* playing a notable role. These findings highlight that, while some microbial interactions are conserved across biomes, most remain highly specialised. Motivated by this distinctness, we next sought to explore the functional implications of these interactions using metabolic modelling, aiming to uncover the potential ecological roles of these community structures.

### Environment-specific metabolite exchange patterns shape microbial community behaviour

To investigate metabolic interactions within microbial communities, we constructed genome-scale metabolic models (GEMs) for all nodes in the co-occurrence networks derived from hospital, metro and office microbiome (Supplementary Table 2). Co-occurrence networks were utilised to define community structures, with community size kept constant across biomes to enable comparisons. All feasible combinations of community sizes ranging from 2–9 members were explored. We then assessed the Metabolic Interaction Potential (MIP) and Metabolic Resource Overlap (MRO) within these communities to understand their metabolic trade-offs (see Methods). MIP quantifies the actual number of metabolites that could be exchanged between members of a community, which is a proxy measure of cooperative interaction. Conversely, MRO indicates the degree of overlap in resource utilisation, which acts as a proxy for competition for shared nutrients. Notably, the office communities exhibited high resource overlap and low interaction potential, indicating a competitive metabolic landscape. On the other hand, hospital and metro communities showed a complementary structure with high interaction potential and low resource overlap. With an increase in community size, the metabolic disparities between biomes grew more pronounced, underscoring the ecological importance of higher-order interactions in community dynamics (Supplementary Figure 4). We therefore focused on the maximal community size (nine members) for subsequent analyses (Supplementary Table 2). PERMANOVA analysis revealed significant divergence in interaction profiles between environments (pseudo-F = 14.49, *p <* 0.001), with the environment explaining 44.3% of the total variance in this ecological interaction space (Figure 5a).

**Figure 5:**
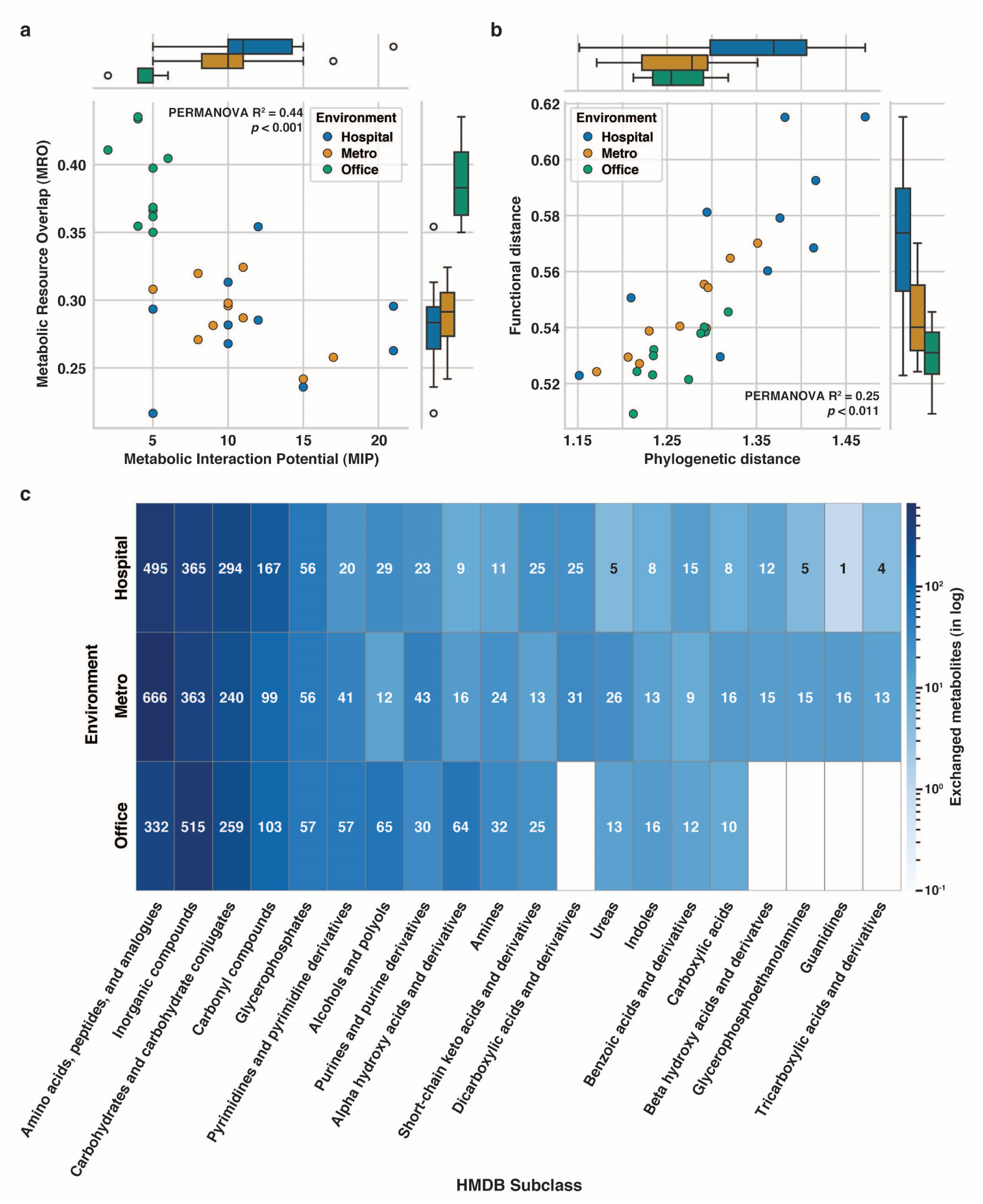
Metabolic cross-feeding among microbial communities in built environments. **a,** Scatterplot of metabolic interaction potential (MIP) versus metabolic resource overlap (MRO) for 30 microbial communities (10 each from hospital, metro and office environments), with marginal box plots showing the distribution of MIP and MRO by environment. Differences across environments were assessed using PERMANOVA based on Bray–Curtis distance (pseudo-F = 14.49, *R*^2^ = 0.44, *p* = 0.001). **b,** Scatterplot of mean phylogenetic distance versus mean functional distance for the same 30 communities, with marginal box plots shown by environment. PERMANOVA based on Bray–Curtis distance indicated significant separation between groups (pseudo-F = 5.12, *R*^2^ = 0.25, *p* = 0.01). **c,** Heatmap showing the top 20 most exchanged metabolite subclasses across the three environments. Values represent the total number of inferred exchanges across 10 communities per environment. Metabolite subclasses are annotated using Human Metabolome Database (HMDB) classifications. The colour scale reflects log-transformed values of total exchanges.

To elucidate the underlying ecological and evolutionary drivers of these patterns, we quantified both phylogenetic and functional distances between community members (Supplementary Table 3). Phylogenetic relatedness was estimated using both alignment-based (marker-gene-based) and alignment-free (Mash) methods, while functional distances were derived from the Clusters of Orthologous Genes (COGs) and metabolic reaction comparisons. Across all communities, phylogenetic distance was strongly correlated with functional distance (Pearson R = 0.84; Figure 5b). Office communities comprised closely related and functionally redundant taxa, consistent with competitive dynamics and high MRO. In contrast, hospital communities were phylogenetically and functionally diverse, supporting the emergence of metabolic complementarity. Metro station communities exhibited an intermediate profile. A PERMANOVA test further confirmed significant environmental contribution (pseudo-F = 5.12, *p* = 0.015), with 25.4% of the variance in functional–phylogenetic space explained by it. Similar trends were observed when comparing phylogenetic distances with metabolic and Mash distances (Supplementary Figure 5, Supplementary Table 3).

To further explore the molecular bases that could explain divergent metabolic profiles in diverse environments, we studied the specific metabolites predicted to be exchanged in nine-member communities across biomes. Metabolites were categorised into broader groups of chemicals according to their HMDB subclass annotations (see Methods, Supplementary Table 2). We observed a combination of conserved and environment-specific patterns of metabolite exchanges (Figure 5c). Across all environments, the most commonly exchanged metabolites included amino acids, peptides and analogues, inorganic compounds, carbohydrates and carbohydrate conjugates. Although commonly exchanged, when compared to other community types, office communities evidently exchanged fewer amino acid derivatives but more inorganic compounds. The hospitals had higher exchanges of benzoic acids and derivatives and carbonyl compounds, which may indicate antibiotic-related selection due to the clinical setting. Interestingly, nucleobase derivatives (pyrimidines and purines) were exchanged less frequently in hospital communities compared to their metro and office counterparts. Moreover, the metro-associated communities showed a distinct enrichment in the exchange of *β*-hydroxy acids, glycerophosphoethanolamines, guanidines, and tricarboxylic acid derivatives, suggesting a possible adaptation to stress from desiccation or fluctuating availability of nutrients. In office settings, the presence of amines, alpha-hydroxy acids, alcohols, and polyols implies fermentation-based metabolism, which potentially could be influenced by high overlap of resource utilisation and stress-resilience mechanisms. Together, these findings demonstrate that while a core set of metabolites dominates microbial interaction in built environments, the nature of these metabolic exchanges is highly context-dependent. Each environment imparts various cooperation-competition balances and metabolic repertoires, showing how microbial communities evolve adaptively, responding to their surrounding habitat.

### Metabolic cross-feeding reveals ecological facilitation of pathogens in hospital environments

In order to investigate the prevalence and ecological support of pathogens within built environment microbial communities, we mapped all identified taxa to the gcPathogen database to determine their pathogenic potential and corresponding biosafety levels (BSL) (Supplementary Table 2). The percentage of pathogens abundant in the three environments was quantified (Figure 6a). As expected, hospital environments had the highest proportion of pathogens (55.9%), demonstrating their ability to serve as reservoirs for a range of opportunistic and nosocomial organisms. Notably, metro (41.8%) and office (40%) environments also exhibited pathogens. When looking at biosafety levels of these pathogens, BSL-2 pathogens dominated all environments, organisms capable of causing mild to moderate diseases in humans. Interestingly, BSL-3 pathogens were detected in both hospital and metro environments but were absent in office settings. This highlights the potential public health implications of microbe transmission throughout urban transit networks, which could serve as a silent conduit for clinically relevant infections. Furthermore, we calculated the Pathogen Support Index (PSI) for each community (Figure 6b) to determine to what extent different environments support pathogenic taxa. PSI represents the level of metabolic support that pathogen species may have as a part of the microbial community. Communities associated with hospitals exhibited significantly higher PSI values than those of metro and office environments, suggesting that hospital microbiomes offer more conducive ecological niches for the persistence and activity of pathogens. For the metro and office communities, the PSI values did not differ significantly, suggesting a comparable, albeit lower, capacity for hosting pathogens.

**Figure 6:**
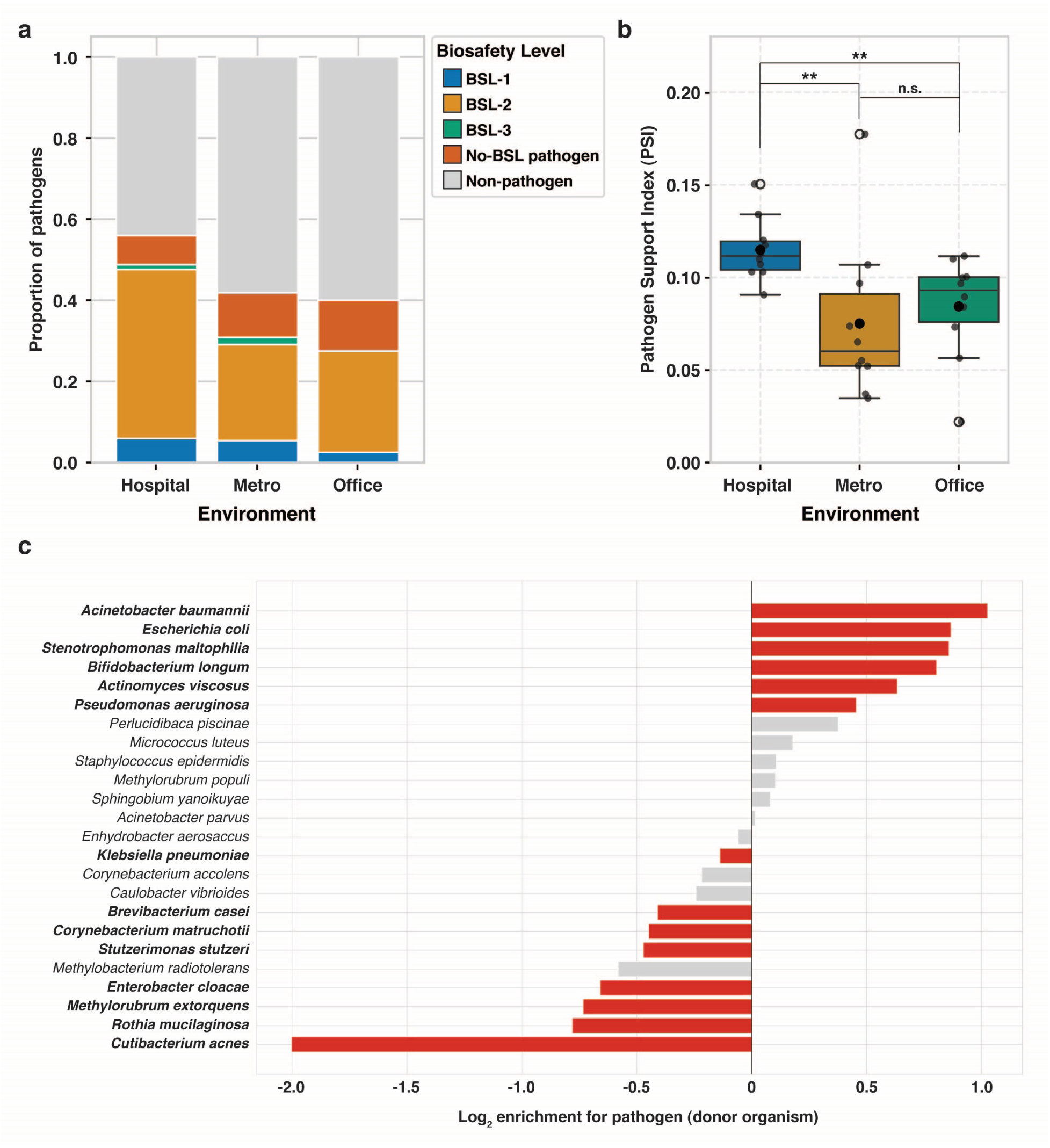
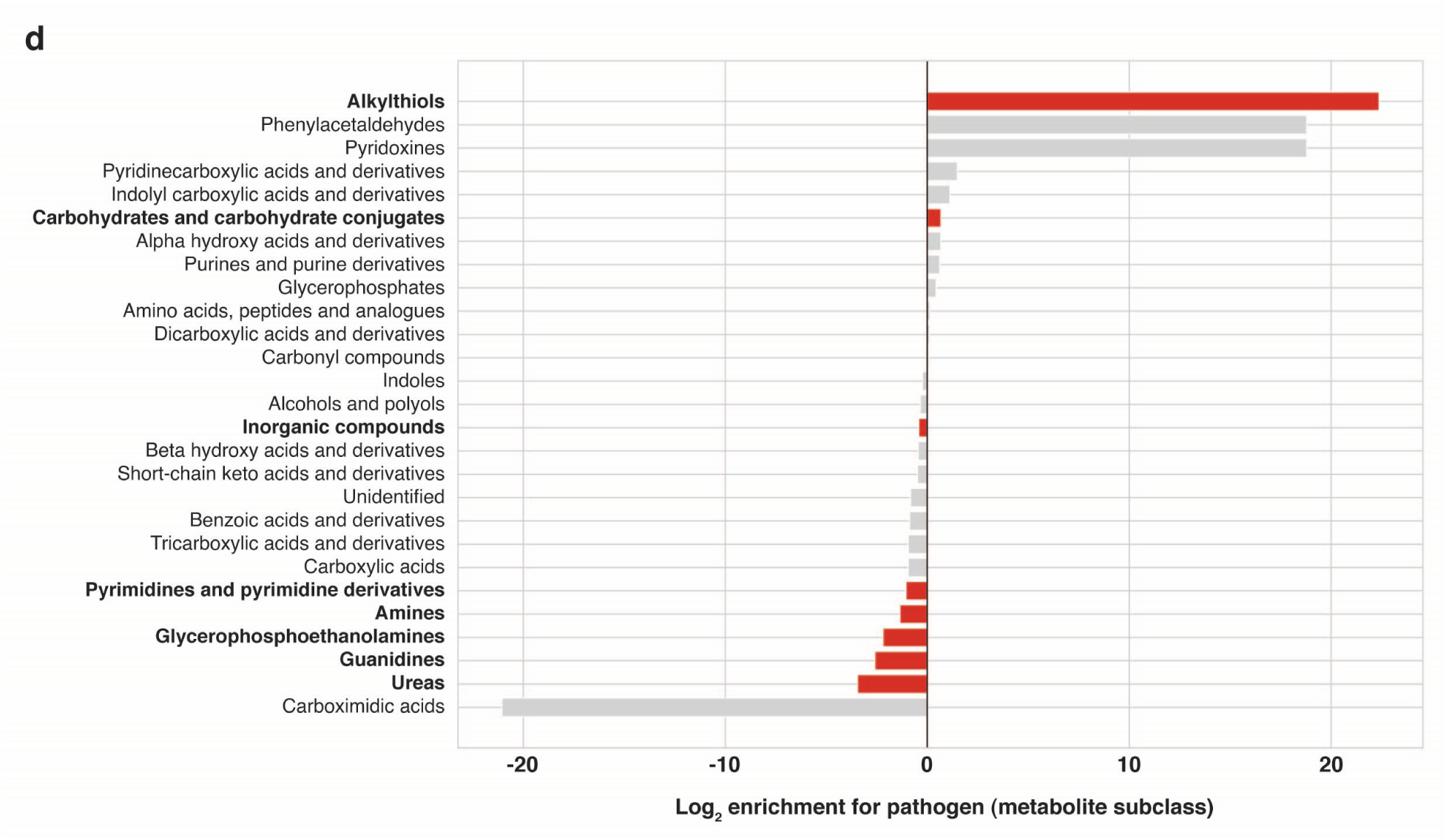
Community-level support for pathogen colonisation across built environments. **a,** Stacked bar plots showing the relative proportions of pathogens across hospital, metro, and office environments. Pathogens are classified by biosafety level (BSL), alongside non-BSL pathogens and non-pathogenic taxa. **b,** Box-and-whisker plots showing the distribution of the Pathogen Support Index (PSI) across microbial communities (*n* = 10 per environment). PSI quantifies the normalised metabolic support provided to pathogens by each community [See ‘Methods’ section]. Significance levels: ** 10^−3^ *< p* ≤ 10^−2^, n.s.: > 0.05. Boxes indicate the interquartile range (IQR), with the central line denoting the median and whiskers extending to 1.5× IQR. The black dot indicates the mean PSI for each environment. **c-d,** Bidirectional bar plots displaying significant cross-feeding interactions involving pathogens and non-pathogens across all environments. **c,** Significant donor species contributing to metabolite exchange with pathogen and non-pathogen groups. **d,** Metabolites significantly exchanged and received by the pathogen and non-pathogen groups. In both panels, the left (blue) bars represent support for non-pathogens, and the right (red) bars represent support for pathogens. Support values represent the aggregate contribution across communities (*n* = 30). Colored bars indicate statistically significant contributors, while non-significant donors or metabolites are shown in grey.

To delve into these pathogen supports, we studied metabolite exchange patterns involving pathogens and non-pathogens in microbial communities. We first profiled donor organisms and assessed the nature of exchanged compounds (Supplementary Figure 6). *Acinetobacter baumannii*, *Escherichia coli*, *Pseudomonas aeruginosa*, *Pseudomonas putida*, *Ruminococcus torques* and *Stenotrophomonas maltophilia* are among the few organisms that have been identified as the bacteria that support pathogen growth using frequent and directed metabolite exchanges (Figure 6c). Their role in metabolically supporting pathogens indicates that they may assist in the survival or virulence of co-occurring pathogenic strains. On the other hand, organisms such as *Brevibacterium casei*, *Cutibacterium acnes*, *Enterobacter cloacae* and *Klebsiella pneumoniae* showed significantly more metabolite exchange with non-pathogens, suggesting that some of them may be opportunistic pathogens themselves but would not have major roles in supporting other pathogenic taxa in a community, perhaps because of competitive exclusion or niche specialisation. At the metabolite level, we observed that carbohydrates and carbohydrate conjugates and alkylthiols were significantly enriched in metabolite exchanges involving pathogens (Figure 6d), while compounds such as inorganic salts, pyrimidines and pyrimidine derivatives, amines and urea were exchanged more frequently among non-pathogenic taxa. Together, these insights shed light not only on the widespread nature of pathogens within built environments, but also on the different ecological contexts that modulate their survival and growth. The presence of key metabolic donors and specific compound classes differentiates the underlying biochemical frameworks that allow pathogens to thrive within complex environmental microbiomes.

## Discussion

In this study, we investigated microbial communities in hospital environments to decipher patterns in microbial diversity, interactions, and metabolic factors that affect community structure and pathogen persistence. Here, our objective was not simply to catalogue microbial diversity, but to gain a metabolic perspective on how these communities assemble and are structured. Previous studies have reported that hospitals represent unique ecological niches that host diverse microbial communities different from those of other built environments [26, 27]. Despite thorough infection control procedures for pathogenic microorganisms [28, 29], microbial contaminations and suppression of opportunistic pathogens remain challenging. Our analysis also indicated an enrichment of Bacilli, Clostridia and Gammaproteobacteria classes, well known for being associated with nosocomial infections and for hosting clinically relevant genera such as *Clostridium*, *Enterococcus*, *Pseudomonas*, and *Staphylococcus* [30]. In addition, we identified a notable fraction of the virome, which constitutes nearly 10% of the total microbial abundance in hospitals. Although viruses in built environments present serious public health risks, they have been vastly understudied compared to bacteria [31]. Given the habitat-specific diversity and metabolic potential of viromes and the emerging evidence for virus-host coevolution [32], understanding their role in microbial dynamics and infection control in healthcare settings should be a priority.

To better understand microbial associations in the built environments, we investigated patterns in the microbial co-occurrence networks. Our analysis showed that hospital networks are more complex in both composition and interactions compared to metro and office networks. This would suggest dynamic heterogeneity in a hospital setting caused by a variety of microhabitats, frequent human activity, and antimicrobial treatments [33]. A similar correlation between microbial richness and network complexity has previously been reported in soil ecosystems, where more diverse habitats produced denser microbial networks with high connections [34, 35]. Furthermore, hospital and office modules were observed to overlap with metro modules, suggesting that transport systems could potentially serve as ecological conduits that bridge communities between static built environments [36]. When we explored hubs in these networks, the hospital network hubs were found to consist of well-characterised nosocomial pathogens such as *Klebsiella pneumoniae* [37], *Staphylococcus aureus* [38], *S. epidermidis* [39], *S. capitis* [40], and *Serratia marcescens* [41], which have historically been able to persist against disinfection, antibiotic therapy, and nutrient limitation. Given their high centrality in the network, these organisms can play a key role in spreading antibiotic resistance and therefore promoting the persistence of the pathogenic community [42]. However, the presence of *Bifidobacterium longum*, a commensal and intestinal probiotic, as a hospital hub may imply elevated use of probiotics, particularly in immunosuppressed patients, and supports its emerging status as a protective biomarker against infections [43]. In addition, the office network hubs were primarily composed of organisms of environmental and commensal taxa, such as *Acinetobacter parvus*, *Microbacterium laevaniformans*, *Methylorubrum populi* and *Mycolicibacterium fortuitum* — organisms that are well adapted for indoor environments due to their desiccation tolerance and ability to survive on airborne and surface-derived substrates for metabolism [44, 45, 46]. Likewise, metro station network hubs such as *Acinetobacter johnsonii*, *Chroococcidiopsis thermalis*, and *Deinococcus wulumuqiensis* were also noted for their tolerance to stress in the variable and high-contact transportation environment [47].

Although the identification of hub species provides critical insight into the dominant species, a comprehensive understanding of microbial ecology requires the analysis of the interactions that shape the community. Our analysis of network edges revealed that Actinobacteria accounted for the highest number of interactions in all three environments, highlighting their ecological ubiquity. Owing to their extreme metabolic plasticity and the ability to synthesise a wide array of secondary metabolites, Actinobacteria may serve as keystone taxa that define the structure and function of microbial communities in built environments [48]. The relatively low fraction of generalised interactions (4.1–23.3%) further supports the fact that microbial associations are highly context-dependent. Our analysis showed that generalised interactions predominantly occurred within members of the same taxonomic classes, while specialised interactions more frequently involved distantly related groups. This indicates that intra-class interactions might be based on conserved functional traits shared among related taxa [49], whereas inter-class specialised interactions may form through environment-specific selective pressures and consequently establish novel functional collaborations between organisms that are phylogenetically distant from each other [50]. However, co-occurrence network-based approaches remain incapable of providing evidence of true biotic interactions [20]. Addressing this gap, we incorporated genome-scale metabolic modelling in these networks to reveal the molecular basis for complex microbial interactions and to explore their implications for ecosystem resilience and pathogenicity.

While our study provides new insights into the microbial ecology of built environments, certain limitations need to be acknowledged. In this study, we utilised the data from Chng *et al*. [6], a benchmark study for the cartography of the hospital microbiome using large-scale shotgun metagenomics, which offers deeper taxonomic and functional resolution than amplicon sequencing commonly used in previous studies. Samples from each environment were also processed in a similar fashion, reducing the possibility of batch effects and false positives. However, despite these advantages, hospital samples studied from a single institution may not sufficiently represent the diverse hospital environments worldwide. Furthermore, our analysis did not explore abiotic factors such as humidity, temperature, and cleaning frequency [51]. Another critical limitation could be the comparatively smaller sample sizes for office and metro environments, compared to hospitals. Recent studies indicate that in order to obtain enough statistical power for microbiome network analysis, substantially larger sample numbers are needed, with a minimum threshold of 25-30 samples per group [52]. Even though the numbers were within the bare minimum needed for a microbiome study, imbalance can introduce bias into comparisons at the network level. To address this, a rarefaction analysis was performed as an estimate of species accumulation between environments so that the microbial richness could be fairly compared [53]. Our bootstrap-based validation of network inferences with an adaptive Benjamini-Hochberg correction was further added to strengthen the robustness of the method. In addition, the compositional nature of microbiome data introduces inherent analytical challenges. We concede the possibility of false-positive and false-negative associations, even with our strict filtering parameters of *>* 1% relative abundance and *>* 10% prevalence [54]. We strove to overcome this by implementing some of the best practices in compositional data analysis, as detailed in the Methods section. The Majorization-Minimization approach in the gcoda was chosen for its effectiveness in handling compositional data and capturing direct interactions in microbial communities [55].

While inferring modelling-based metabolite exchanges, SMETANA considered a number of assumptions. It is assumed that all community members grow simultaneously, without any temporal dynamics, spatial heterogeneity, or environmental constraints. Gap-filling in genome-scale metabolic model reconstruction was purposely omitted to avoid false-positive predictions; hence, the inference could underestimate true metabolic capabilities [20]. However, in spite of such shortcomings, the model approach presented still offers mechanistic evidence for metabolic exchanges demonstrated in many previous studies spanning diverse environments [20, 56, 57]. The usage of type strains and representative genomes from the NCBI RefSeq database ensures taxonomic accuracy, whereas the use of multiple distance metrics (phylogenetic, functional, and metabolic) improves the robustness of the given community structure analyses. Our metabolic model-based studies showed that different built environments foster different patterns of metabolic cooperation and competition. This has been demonstrated by contrasting Metabolic Interaction Potential (MIP) and Metabolic Resource Overlap (MRO) profiles [16]. Given their complex nature, offices may pose as a resource-limiting environment and thus reduce microbial diversity and favour competition among functionally similar taxa [58]. In contrast, the hospitals and metro communities exhibited complementary metabolic structures with high interaction potential and low resource overlap, supporting the hypothesis that environmental complexity drives cooperative metabolic interactions [16, 59]. In addition, the strong correlation between phylogenetic and functional distances found across communities emphasises the fundamental principle of microbial ecology where evolutionary relatedness constrains functional capabilities [60, 61]. Identification of amino acids, peptides, inorganic compounds, and carbohydrates as the most commonly exchanged metabolites in all environments signifies the core aspects of microbial metabolism and cross-feeding [62]. Amino acids are possibly most extensively described as key currency molecules amongst microbial communities, with usages for protein biosynthesis or energy metabolism [63]. Moreover, environment-specific metabolic signatures were evident as well. These findings highlight that while core metabolic exchanges do support microbial interactions in built environments, specific metabolite exchange patterns are highly context-dependent and could serve as biomarkers for microbiome health and inform targeted strategies to promote beneficial microbial communities.

Lastly, we were interested in understanding whether some communities support pathogenic organisms. While our study focuses primarily on hospital environments, it is important to contextualise the presence of pathogens within the broader urban microbiome. We observed that pathogens are ubiquitous and not exclusive to hospitals. However, the presence of pathogens across urban built environments should not obscure the critical qualitative and mechanistic differences that distinguish hospitals from other enviroments. Estimation of the Pathogen Support Index (PSI) thus provides a novel way to quantify the actual metabolic support that contributes to pathogen persistence in environmental microbiomes. The significantly higher PSI values observed in hospital communities are consistent with research that demonstrates that hospital environments create unique ecological niches that promote pathogen survival and persistence despite routine disinfection practices [64]. Identification of microbial taxa that provided metabolic support to pathogenic organisms allows us to understand the underlying ecological mechanisms behind the facilitation of pathogens in built environments. Recent research showed that ESKAPE pathogens such as *Klebsiella pneumonia* and *Enterobacter bugandensis* obtain complex metabolic support from their neighbouring microbial communities aboard the International Space Station, a unique space-based built environment [65, 66]. Another ESKAPE pathogen, *Pseudomonas aeruginosa*, has also been shown to have complex lactate metabolism, allowing metabolic cross-feeding interactions through the use of different lactate enantiomers [67]. The degradation of complex carbohydrate structures and the release of nutrients for other members of the community can potentially place the organism as an important facilitator of pathogen growth [68]. Moreover, the differential enrichment of metabolites between the pathogenic and non-pathogenic groups indicates fundamental differences in metabolic strategies among pathogenic and commensal organisms. Identifying specific metabolic donor organisms and their associated compound classes could act as potential candidate targets for disrupting the pathogen support network as well as infection control strategies in built environments [2, 69]. Understanding the metabolic dependencies of pathogenic organisms opens possibilities for disrupting pathogen support networks through targeted interventions that modulate specific metabolite exchanges.

## Conclusion

Using a comprehensive approach, this study investigates the taxonomic and metabolic interdependencies of microbial communities in built environments, with a focus on hospitals. The combination of genome-scale metabolic modelling and network analysis provides a promising strategy to further understand microbial interactions and inform public health efforts in mitigating microbial risks in urban indoor spaces. Through our unique Pathogen Support Index, we uncover intricate ecological dynamics that promote pathogen persistence in hospital settings compared to other environments. Identifying which metabolites are exchanged and the corresponding donors reveals the metabolic underpinning of pathogen ecology in built environments. These findings highlight the need to incorporate ecological and metabolic contexts in designing effective urban microbial management strategies.

## Data availability

All of the data required to generate the results and figures presented in this article are publicly available in the following repository: https://github.com/RamanLab/hospital_microbiome

## Code availability

All of the codes required to generate the results and figures presented in this article are publicly available in the following repository: https://github.com/RamanLab/hospital_microbiome

## Acknowledgements

P.S. is a recipient of the Prime Minister’s Research Fellowship (PMRF) from the Ministry of Education, Government of India. The authors thank Dr. Narendra Dixit, Dr. Maziya Ibrahim, and Dr. Aarti Ravindran for their valuable insights on this manuscript.

## Author Contributions

Conceptualisation: P.S. and K.R. Methodology: P.S. and V.S.K. Investigation: P.S. and K.R. Supervision and funding acquisition: K.R. Writing—original draft: P.S. Writing—review and editing: P.S., V.S.K., and K.R.

## Funding

P.S. is supported through the Prime Minister’s Research Fellowship from the Ministry of Education, Government of India. K.R. acknowledges support from the Wadhwani School of Data Science and AI, Indian Institute of Technology Madras. Sponsors had no role in study design, data collection and interpretation, manuscript writing, or decision to submit the work for publication.

## Competing interests

The authors have no conflicts of interest to declare that are relevant to the content of this article.

## Methods

### Data

Microbial abundances associated with 438 hospital samples from Tan Tock Seng Hospital (TTSH) in Singapore were previously quantified through shotgun metagenomic sequencing [6]. Along with these samples, we incorporated metagenomic data from metro stations (*n* = 99, [25]) and an office environment (*n* = 30, [6]) within Singapore to enable a comparative analysis of microbial interactions in diverse built environments. Samples lacking metadata information (*n* = 10) were excluded from subsequent analyses. To elucidate the taxonomic composition of the identified microorganisms, we employed the Python toolkit ete3 v.3.1.3 [70] and retrieved lineage information from the NCBI Taxonomy database. We conducted rarefaction analysis to estimate species accumulation across environments, enabling comparison of microbial richness at varying sample sizes. Microorganisms were then categorised into four major taxonomic groups: Archaea, Bacteria, Fungi, and Viruses. However, our focus is only on bacteria, given their predominant representation in the microbial population.

### Statistical analysis of bacterial composition

To systematically explore microbial composition across the environments, we converted the microbial relative abundance data, taxonomy information, and metadata into a phyloseq object using R v.4.3.2. Utilising the microbiome package (https://github.com/microbiome/microbiome/) in R, we calculated microbial alpha diversity using Shannon index metric, a metric that considers both species richness and evenness, representing the diversity of microorganisms within individual samples. The nonparametric Kolmogorov–Smirnov test was employed to determine the significance of alpha diversities across all-vs-all environments, and the *p*-values were adjusted using the Benjamini–Hochberg method. We extended our analysis by randomly selecting an equal number of samples from each environment and computed Shannon diversity across 100 iterations to evaluate alpha diversity across varying sample sizes. Further, we performed beta diversity analysis using Bray–Curtis distance matrices and applied nonmetric multidimensional scaling (NMDS) for ordination. The impact of various factors on community dissimilarity was tested using PERMANOVA (Permutational Multivariate Analysis of Variance) with randomisation (*n* = 999) through the adonis2 function from the vegan package in R, based on Bray-Curtis distances. The significance was measured using pseudo-F statistics. Bacterial composition, with a minimum detection threshold of 0.001 and 10% prevalence, was plotted after agglomeration at the class level.

### Co-occurrence network construction

We focused on potential keystone species by selecting bacteria present in at least 10% of samples with a relative abundance of *>* 1% in at least one of them. Subsequently, we excluded unclassified bacteria and retained only microorganisms demonstrating substantial abundance and prevalence within the particular environment for interaction analyses. We employed the NetCoMi v.1.1.0 (Network Comparison for Microbiome data) in R to investigate microbial interactions within built environments, leveraging individual phyloseq objects derived from diversity analysis [71]. Recognising the compositional nature of the data, we implemented several strategies for constructing the microbial network. Initially, zero values were handled by incorporating pseudo counts using a non-parametric function called multRepl. This function introduced small values (*δ*) for all zeros and adjusted non-zero values while adhering to a unit-sum constraint to preserve their covariance structure.

Subsequently, we normalised the abundances using a Total Sum Scaling (TSS) transformation, and the normalised abundances served as the basis for constructing the microbial network using the gcoda method, a conditional dependence method that effectively captures direct interactions in microbial communities. The gcoda algorithm employs a Majorization-Minimization approach to infer the covariance matrix from the data, maximising a penalised likelihood, particularly effective for compositional data [55]. A ‘signed’ method was employed to manage negative associations, transforming sparsified associations into dissimilarities. A quantitative network assessment was performed through a permutation approach (1000 bootstraps) with an adaptive Benjamini–Hochberg correction for multiple testing (alpha = 0.05). The resulting inferred similarities were utilised as edge weights in the constructed microbial network.

### Network analysis and visualisation

We utilised the netAnalyze function within NetCoMi to evaluate the key topological characteristics of the network, with a focus on the largest connected component. Then, we applied the cluster_fast_greedy, a fast greedy modularity optimisation method from the igraph package to identify dense subgraphs within different environments. These subgraphs were then hierarchically clustered using Jaccard distance based on taxonomic presence. To further understand the network, we calculated normalised centrality metrics such as degree, closeness centrality, and eigenvector centrality. The top 10 nodes with the highest normalised centrality scores were identified as network hubs. For network visualisation, we employed the plot.microNetProps function, leveraging igraph v.1.4.2 and utilising the Spring layout algorithm to generate graphics. Node size was adjusted based on the normalised degree, and each subcluster within the network was assigned a distinct colour. The unconnected nodes were removed.

Next, we distinguished the generalised and specialised interactions by examining edges and categorising them into four major groups. Edges present in multiple networks were considered generalised edges, while those specific to one network were deemed specialised edges. Similarly, a node present in multiple networks is classified as a generalised node, whereas a node found exclusively in a single network is designated as a specialised node. We then classified specialised edges into three subgroups: those with both unique nodes for the environment (specialised edges with both specialised nodes), those with only one unique node for the environment (specialised edges with one specialised node), and those with both nodes present in multiple environments (specialised edges with generalised nodes). We then evaluated the distribution of these categories across the environments.

### Genome retrieval, annotation and reconstruction of genome-scale metabolic models

In order to assess the metabolic interactions within the microbial communities, the genomes of the identified microorganisms were mapped from the NCBI Reference Sequence (RefSeq) database (accessed in May 2024). We specifically obtained type strains of the organisms by selecting ‘assembly from type material’, ‘assembly from synonym type material’, or ‘assembly designated as neotype’. In the absence of type strains, the NCBI representative genome or the available genome was collected. The genomes were then downloaded in FASTA format using the command line tool bit v.1.9.4 [72]. We annotated the acquired genomes using Prokka v.1.12, which utilises Prodigal for gene identification and annotation [73].

Genome-scale metabolic models (GEMs) were then reconstructed for all identified organisms using their translated CDS sequences in protein FASTA format with CarveMe v.1.6.0 [74]. CarveMe was run with specific curated templates for Gram-positive or Gram-negative bacteria, as appropriate, using the IBM CPLEX v.22.10 solver. We intentionally omitted gap-filling to avoid false-positive predictions of cross-feeding interactions. The quality of the models was checked using MEMOTE v.0.17.0 [75]. Growth rates for all models were estimated keeping biomass as the objective function using the optimize function in COBRAPy v.0.29.1 [76].

### Phylogenetic, functional and metabolic distance calculation

We employed both alignment-free and alignment-based approaches to estimate the phylogenetic distance of the identified organisms. First, we used the alignment-free Mash distance approach. Pairwise genomic distances among community members were calculated using Mash v.2.3, with a sketch size of 1000 and a k-mer size of 21 [77]. For the alignment-based method, we used 74 universal marker genes present in bacteria for alignment using Muscle v.5.1 [78]. We utilised GToTree v1.7.07 to construct a phylogenetic tree based on the alignment, and the tree was visualised and annotated using iToL v.6.0.0 [79, 80]. The phylogenetic distances among species were computed using the Biopython Phylo package. We further estimated the functional distance of these microorganisms by clustering their Cluster of Orthologous Genes (COGs). The Prokka annotated fasta amino acid (.faa) files were utilised for the classification of the COGs using COGclassifier v.1.0.5. The presence/absence of the COGs was used to identify the distance between the genomes using Jaccard distance, and the distance is considered as functional distance among the organisms. To assess metabolic distance, we compared the presence/absence of the reactions in the GEMs of the organisms. All the models were read using COBRAPy. Jaccard distances based on reaction presence/absence represented the metabolic distances between genomes.

### Community selection and prediction of cross-feeding potential

Microbial communities were selected from the co-occurrence networks of each environment, with varying community sizes. Here, we defined a community as a connected subgraph of the network with the given community size, with all the members co-occurring at least in one sample. For each environment and community size, we created a set of 10 communities in order to account for the variability in the community composition. SMETANA v.1.2.0 was then employed to simulate microbial communities based on reconstructed GEMs to estimate possible metabolic interactions [16]. SMETANA employs mixed-integer linear programming (MILP) and ensures that all species exhibit non-zero growth under a mass balance constraint. It performs constraint-based analysis for all community members simultaneously without any biological objective function being applied.

We ran SMETANA in both global and detailed modes using the IBM CPLEX v.22.10 solver with flags –flavor bigg –molweight. In global mode, SMETANA assumes a complete growth medium and computes two key interaction scores: i. Metabolic Interaction Potential (MIP), which captures the ability of species to exchange metabolites while reducing their dependence on external metabolites; and ii. Metabolic Resource Overlap (MRO), which determines the degree of competition for shared metabolites among the community members. Subsequently, we ran SMETANA in detailed mode to investigate metabolite exchanges within the community using a community-specific minimal medium. In detailed mode, a pairwise SMETANA score is computed for all the possible interactions by integrating three additional metrics: Metabolite Production Score (MPS), Metabolite Uptake Score (MUS), and Species Coupling Score (SCS). The sum of the SMETANA scores in a given community is considered a community SMETANA score. The mathematical formulation, as well as the description of the different scores, are described in the original publication [16].

Further, the exchanged compounds were classified into broader chemical subclasses by mapping them to the BIGG database (http://bigg.ucsd.edu) and then categorising using the Human Metabolome Database (HMDB; https://www.hmdb.ca/) to assign each metabolite to its subclass (e.g., amino acids and derivatives, carbohydrates and conjugates, etc.). Metabolites that could not be assigned to any subclass were labelled as “Unidentified”.

### Estimating pathogen support through community-level metabolic cross-feeding

To measure the amount of metabolic support that microbial communities provide to pathogenic members, we identified pathogens in each community using the gcPathogen database [81], which provides curated pathogen designations and biosafety level classifications. Pathogen Support Index (PSI), a score derived from SMETANA-predicted metabolite exchange and captures the proportion of total metabolic support directed specifically toward pathogens, was calculated as:

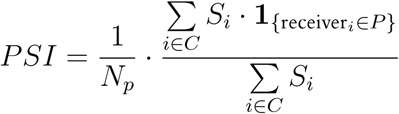

where *C* is the set of all metabolite exchanges in the community, *S_i_* is the SMETANA score of interaction *i*, *P* is the set of pathogens, **1**_{receiver_,_∈P_ } indicates whether the receiver is a pathogen, and *N_p_* is the number of pathogen taxa in the community. A higher PSI value implies increased metabolic assistance toward pathogens in the community.

We further determined which species acted as donors in metabolite exchanges by computing the distribution of compound transfers from these donors towards pathogenic versus non-pathogenic recipients. We accounted for the variation in the overall number of exchanges by normalising the count of transferred compounds by the total number of interactions each recipient group (non-pathogen or pathogen) experienced, reducing biases due to imbalance in the numbers of pathogen and non-pathogen recipients. We then used Fisher’s exact tests with subsequent False Discovery Rate (FDR) correction (Benjamini–Hochberg method) to determine donor species with statistically significant enrichment in their support towards either group. Subsequently, compound-level exchange patterns were analysed to determine if specific metabolites are more frequently transported to pathogens than non-pathogens. These counts were normalised by the total number of recipient-specific interactions to correct for class-wide differences in compound exchange. Subclass enrichment among these chemicals was similarly evaluated using Fisher’s exact tests, with FDR correction.

### Statistical analyses

All analyses were performed in Python v.3.10 and R v.4.3.2. The nonparametric Kolmogorov–Smirnov test was implemented in R through the ks.test function from the stats package v.4.3.1. PERMANOVA (Permutational Multivariate Analysis of Variance) was performed in R using adonis2 function from the vegan package v.2.6.4 and in Python with the permanova function from scikit-bio package v.0.6.3. The Benjamini–Hochberg method was used for multiple testing corrections with the multipletests function from the statsmodels package v.0.14.0 in Python. Unless indicated otherwise, all remaining statistical tests were performed in Python using SciPy v.1.14.0. All data analysis sub-packages were installed in environments using Conda v.24.11.3, and the corresponding yaml files are provided on GitHub.

## Notes

### Competing Interest Statement

The authors have declared no competing interest.

### Funding Statement

P.S. is supported through the Prime Ministers
Research Fellowship from the Ministry of Education, Government of India. K.R. acknowledges support from the Wadhwani School of Data Science and AI, Indian Institute of Technology Madras. Sponsors had no role in study design, data collection and interpretation, manuscript writing, or decision to submit the work for publication.

