## Supplementary Figure for "Microbial Communities Facilitate Pathogen Persistence in Hospital Environments"

#### 13 Supplementary Figures

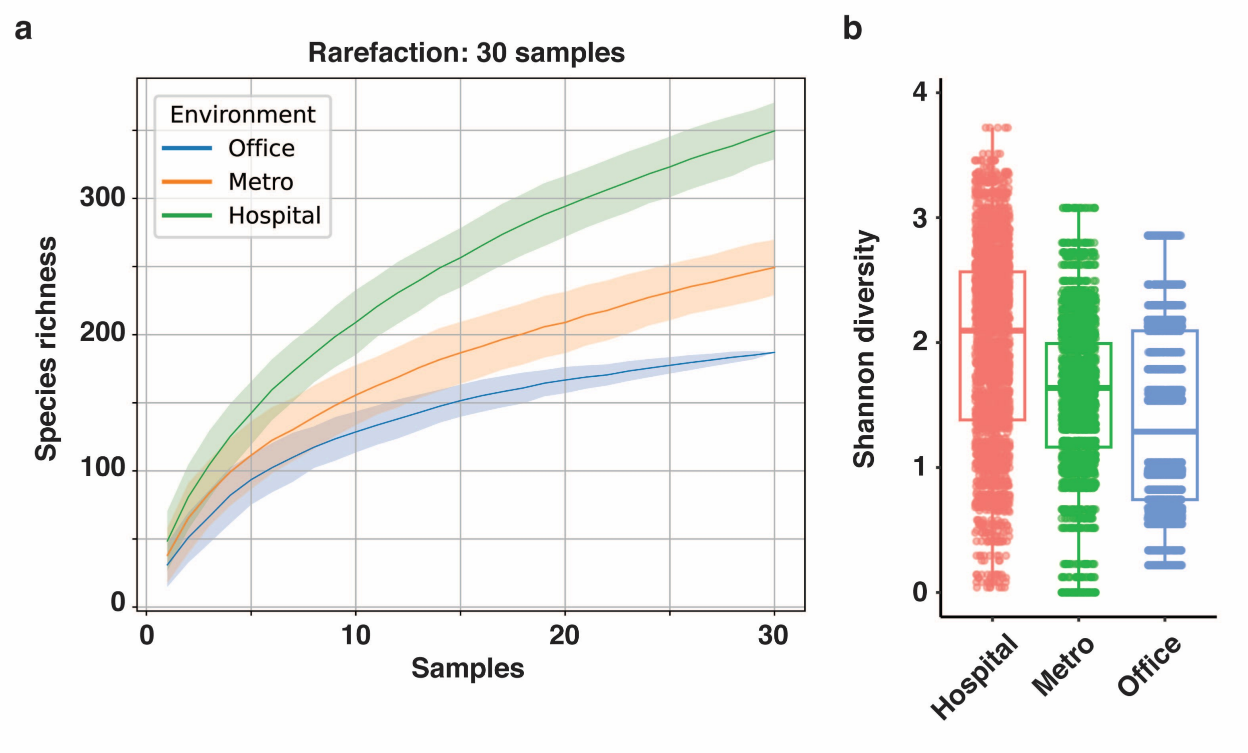


Supplementary Figure 1: **a,** Species accumulation curves demonstrating species richness across environments. The hospital environment exhibited the highest species richness when rarefied to the lowest sample size ($n = 30$) present among the biomes. **b,** Box plots show the distribution of Shannon diversity values for microbial communities in hospital, metro, and office environments, based on 100 iterations of rarefied subsampling, with 30 samples per environment randomly selected for each iteration.

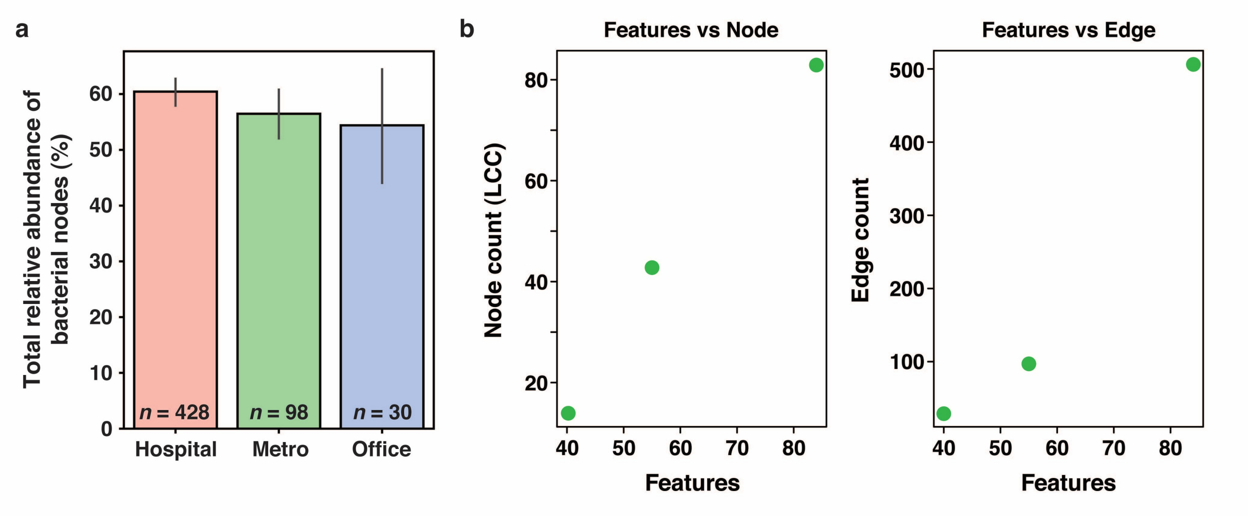


Supplementary Figure 2: **a,** Total relative abundance of bacterial nodes across the three built environments, showing similar levels of microbial diversity with hospital, metro stations, and office environments, all pertaining to approximately 55-60% relative abundance. Error bars represent 95% confidence intervals. **a,** Relationship between the number of features (microbial taxa) and network properties. Left panel: Node count of the largest connected component increases with the number of microbial taxa in the environment. Right panel: Edge count shows a similar increment to the taxa count number.

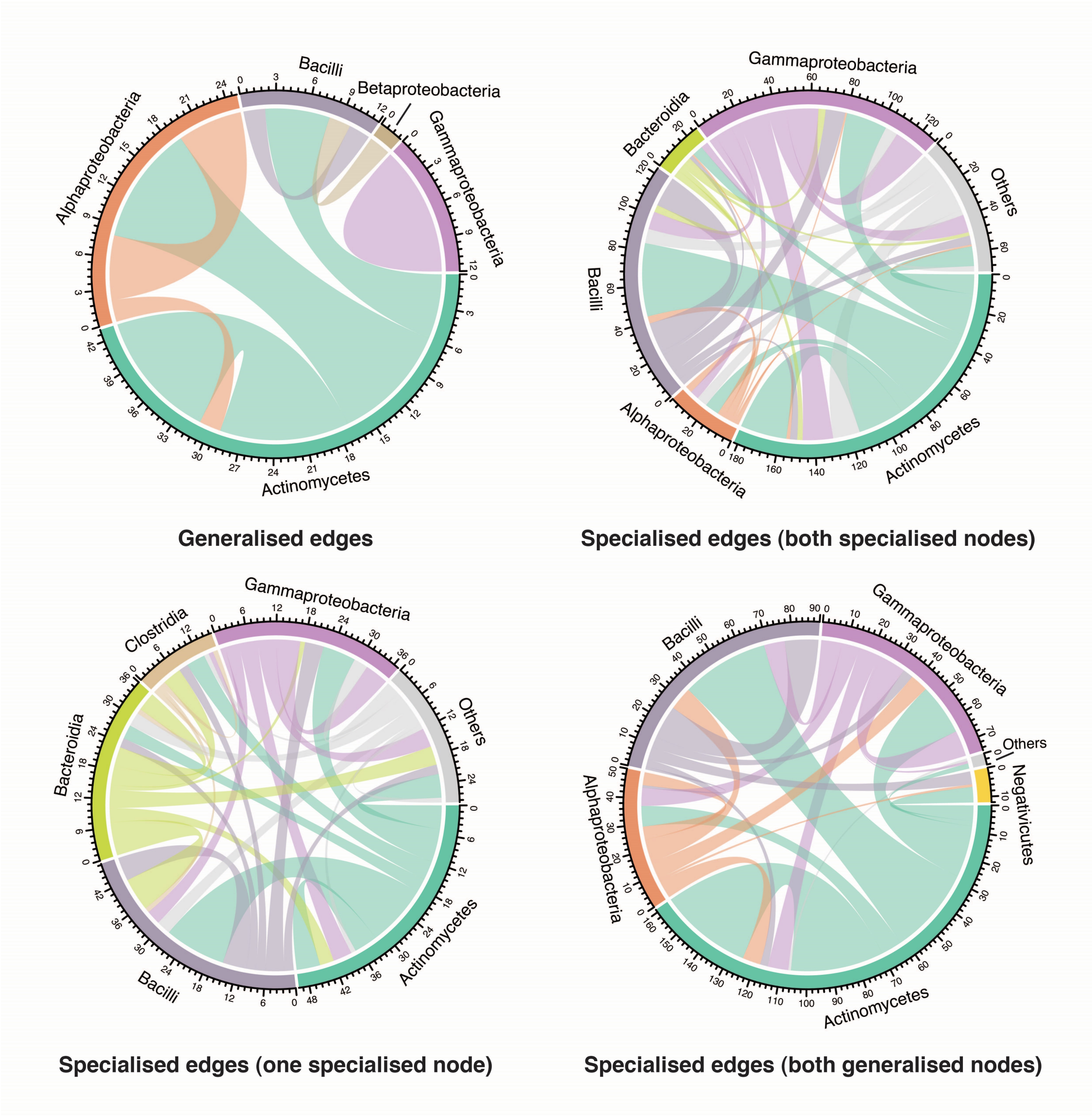


Supplementary Figure 3: Distribution of class-level edges in each of the broad edge categories as shown in Figure 4b.

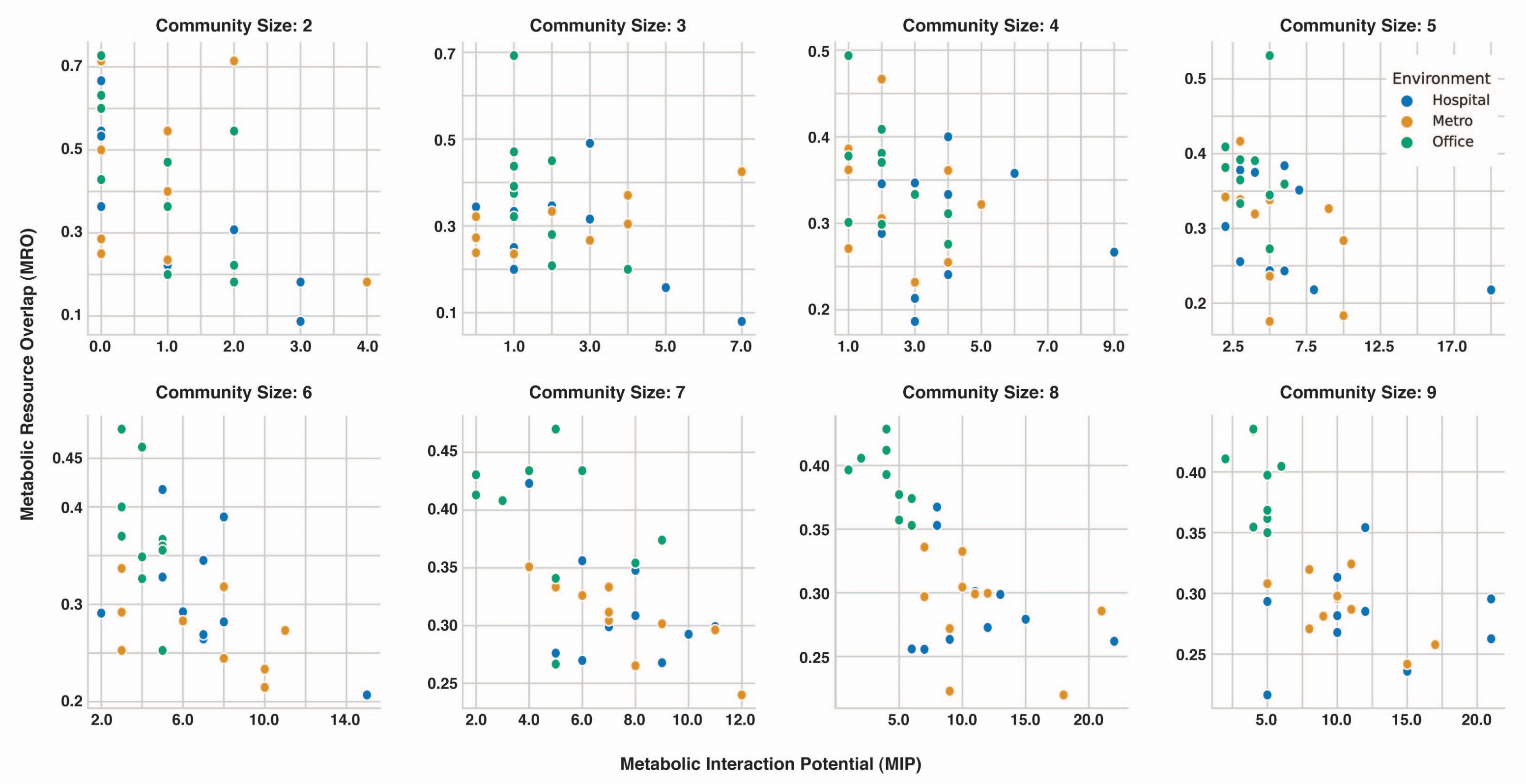


Supplementary Figure 4: Scatter plot showing the distribution of Metabolic Resource Overlap (MRO) and Metabolic Interaction Potential (MIP) across microbial communities from different built environments, for community sizes ranging from 2 to 9 members.

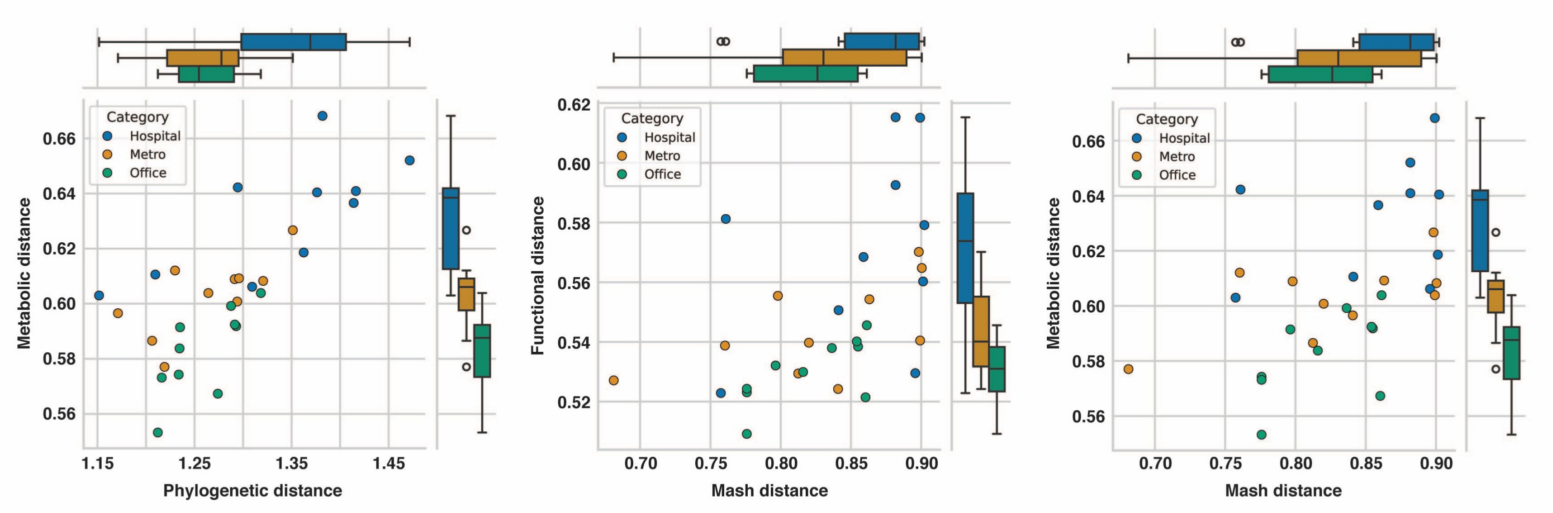


Supplementary Figure 5: Three scatter plots show the correlation between different distance metrics for microbial communities from hospital, metro, and office environments. Marginal box plots on the sides summarise the distribution of distance values for each environment. Individual data points represent the average distance of the community members ($n = 9$) based on either phylogenetic/ functional diversity.

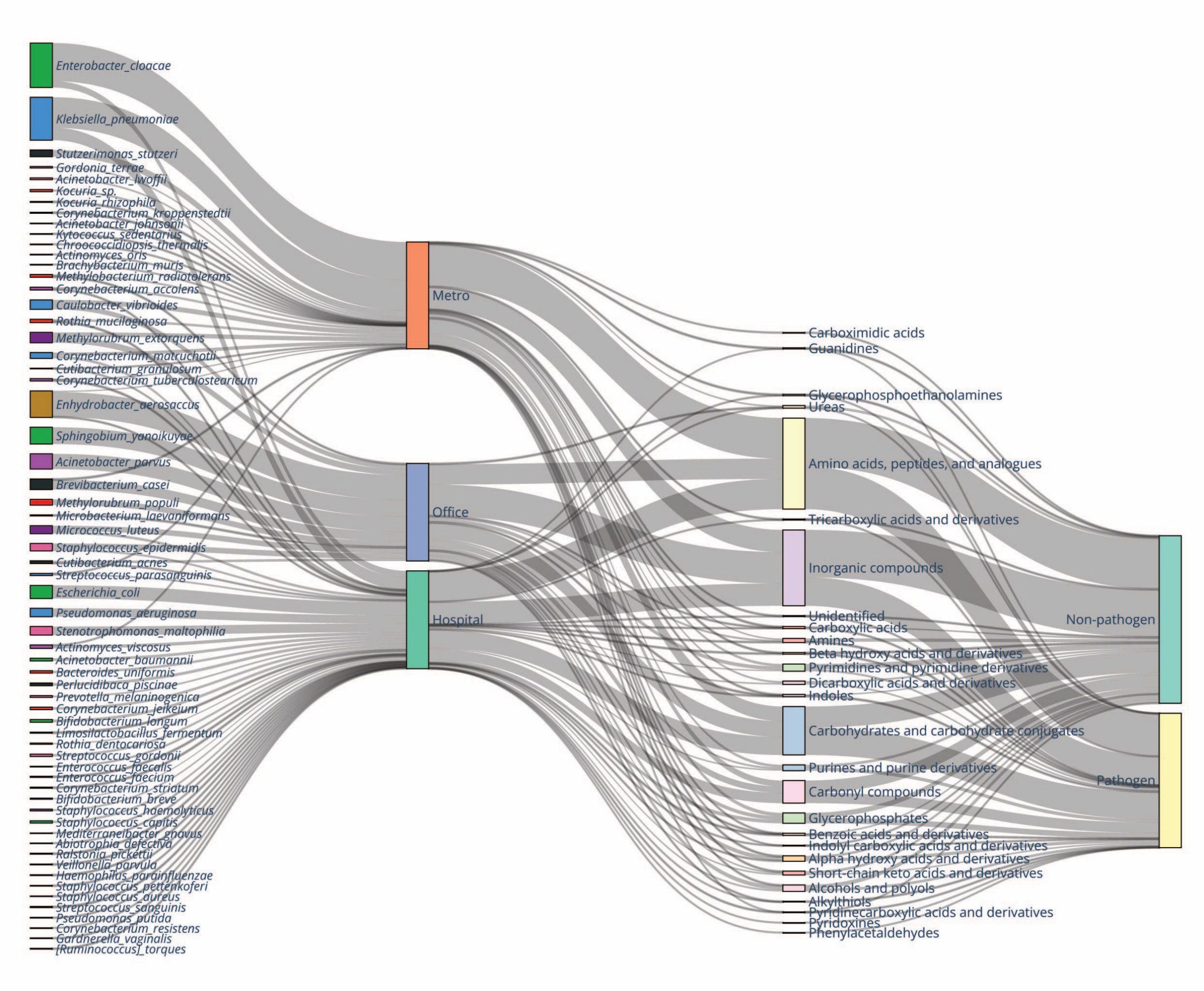


Supplementary Figure 6: Diagram showing the flow of metabolites from donor microbial taxa (through the environment) to the recipient (segregated based on their pathogenic status). The diagram reveals environment-specific patterns of microbial metabolic interactions and their relationship to pathogenic potential across built environments.

### Supplementary Tables

**Supplementary Table T1:** Summary of organism and edge presence–absence patterns, module-wise class counts, and hub information across the urban built environments.

**Supplementary Table T2:** Details of the genome used for metabolic modeling, model statistics, community-structure information, metabolite exchanges, metabolite classification, and pathogen classification of the organisms analysed in this study.

**Supplementary Table T3:** Summary of pairwise distances between organisms based on phylogenetic, functional, metabolic, and Mash distance measures.
